# Improvement of language function after Contralateral Seventh Cervical Nerve Transfer in hemiplegic patients combined with post-stroke aphasia: a prospective observational cohort study

**DOI:** 10.1101/2023.05.18.23290080

**Authors:** Juntao Feng, Tie Li, Minzhi Lv, Miaomiao Xu, Jingrui Yang, Fan Su, Ruiping Hu, Jie Li, Yundong Shen, Wendong Xu

## Abstract

**Background:** While the contralateral seventh cervical nerve (CC7) cross transfer was designed to reconstruct paralyzed arm function after stroke, improvement in language function was found in patients combined with aphasia.

**Objective:** To evaluate the effect of improvement in language function after CC7 cross transfer in stroke patients with chronic aphasia and explore its potential mechanism.

**Methods:** In a prospective observative cohort, patients diagnosed with hemiplegia combined with aphasia were included. The language function was evaluated through the changes of Aphasia Quotient evaluated by Western Aphasia Battery (WAB-AQ) as well as its four subtests from baseline to 1 week and 6 months after the surgery. Patients also received oral agility test by Boston Diagnostic Aphasia Examination (BDAE-OA). Resting-state functional MRI (rs-fMRI) was scanned before and over 6 months after the surgery to explore the potential central mechanism in language improvements.

**Results:** The average increase of WAB-AQ was 8.08 points from baseline to 1 week post-operatively (P<0.001, 95%CI: 5.05-11.10), and 9.51 from baseline to 6-month (P<0.001, 95%CI: 6.75-12.27). In 8 patients who participant in BDAE-OA, the average increase was 3.7 points (95%CI: 0.56-6.84; corrected P =0.023) from baseline to 1-week follow-up, and 5.3 points from baseline to 6 months follow-up. Significant higher local activity was detected at right precentral cortex, right gyrus rectus, and right anterior cingulate cortex after the surgery from rs-fMRI.

**Conclusions:** Immediate and stable improvement in language function was detected after CC7 cross transfer in hemiplegic patients combined with aphasia, which may be realized through enhanced function of language network in the bilateral hemisphere.

## Introduction

Aphasia is a common and disabling sequela after stroke^1^. It has been estimated that more than one third of the stroke patients have symptoms of aphasia of different severity^2^. In patients who enter the chronic phase (generally defined as >6 months or 1 year), slight or none spontaneous progress could be seen^3, 4^. While the speech and language rehabilitation treatment (SLT) has been proved to be effective in improving the symptoms, the effect sizes are often unsatisfactory to demand in daily life ^5-7^. Therefore, more effective and long-term stable interventions are still urgently needed.

As patients with right limb paralysis due to left hemispheric injury are often combined with aphasia, a unique phenomenon is frequently mentioned that, after spinal nerve resection such as selective dorsal rhizotomy, the patient’s language function can be significantly improved^8, 9^. However, this phenomenon has not been fully quantified since the surgical procedure varies in each patient. Moreover, the mechanism underlying this phenomenon is not carefully studied either. Therefore, to what extent spinal nerve resection surgery can benefit language function and through which kind of mechanism still needs to be evaluated.

The contralateral cervical seventh nerve (CC7) cross transfer has been proved to effective in improving the paralyzed upper limb function of chronic hemiplegia patients due to stroke or other brain injuries^10^. In this surgery, one of the five nerve roots of the brachial plexus, the C7 nerve root, is rewired from the contralateral side to the paralyzed side, which established a direct connection between the paralyzed upper limb and its ipsilateral hemisphere, the intact hemisphere. Through animal and clinical observation of the brain plasticity, it shows that this surgery will induce large-scale functional plasticity^11^. In the following observational study, it has been reported that, the functional improvement is not restricted to the upper limb function, but also in gait and lower limb spasticity^12^. Therefore, this surgery seemed to induced systematic functional remodeling rather than improving motor function alone. With a long-term observation of these patients, we found that this surgery can also significantly improve the language function in hemiplegia patients combined with chronic aphasia. This surgery provided a better study model for exploring this phenomenon.

Therefore, in this study, we aim quantify the effectiveness and stability of the improvement in language through a prospective cohort study with six-months follow-up. Since functional MRI has contributed important evidences for understanding of language networks and compensational mechanisms in aphasia patients^1^, we collected the resting-state functional MRI data before and after the surgery^13, 14^. Therefore, this fMRI data can provide preliminary evidence for the potential changes after CC7 nerve cross transfer surgery. Previous studies also indicated the influence of somatosensory and motor signal on the language performance; thus, the motor and spasticity changes were also recorded in this study to evaluate the potential connections between motor and language function.

## Methods

### Study Design and Participants

This is a single center, prospective, observational cohort study approved by the institutional review board. Consent of the study were acquired for all patients participated in this study. In this study, we included patients diagnosed with right hemiplegia combined with aphasia (> 1 year) who were planned to receive contralateral C7 nerve cross transfer surgery to improve the right upper-limb function. The post-stroke aphasia is diagnosed by the Aphasia Quotient (< 93.8) calculated by the Western Aphasia Battery (WAB). The participants should be adults (> 18 years), right-handed, native Chinese speakers with adequate visual and auditory abilities to complete the assessments. Participants were excluded, if they had a speech and language disorder caused by other deficits or before the last disabling stroke (such as neurodegenerative diseases), if they were unable or unwilling to participant in the study, or if they were currently receiving intensive speech and language therapy within four weeks before screening.

### Intervention

#### CC7 cross transfer Surgery

The description of the surgical procedure was reported in our previous study and a surgical technique introduction article^10, 15^. The surgery is main performed in three steps, transection of the proximal C7 nerve root on the paralyzed side, transection of the distal C7 nerve root on the intact side, and transferring the intact C7 nerve root to the paralyzed side and anastomosis with the paralyzed C7 nerve root. Briefly, the bilateral brachial plexus nerves were exposed through supraclavicular incision. The C7 nerve on the paralyzed side is severed near the intervertebral foramen, and the C7 nerve on the intact side is severed as distally as possible before its divisions combine with other divisions from the upper trunk and lower trunk. Then the proximal cut end of the C7 nerve on the intact side is drawn through the prespinal route to the paralyzed side and anastomosed directly to the distal cut end of the C7 nerve on the paralyzed side.

#### Postsurgical Rehabilitation

All patients received standard rehabilitation program according to Huashan Rehab Protocol after CC7 cross transfer surgery^16^, including spasticity control, physical treatment and occupational treatment depending on the nerve regeneration process. During the whole process, no standard speech or language rehabilitation treatment were provided for the patients.

### Outcomes

The language and motor function were evaluated at baseline, and 1-week, 6-month after the surgery. The outcomes were defined as the changes of the tests from baseline to follow-up points after the surgery.

For the language function, the WAB and Boston Diagnostic Aphasia Examination (BDAE) scale were used. The WAB applied in this study is consisted of four subtest, spontaneous speech, auditory comprehension, naming and repetition, with a total score of 20, 10, 10 and 10 points, respectively. Based on these four subtests of the WAB scale, the Aphasia Quotient (AQ) is calculated, with a total score of 100 points. In this study, the oral agility subtest from the BDAE scale was evaluated in patient included after 2021, which is consist of verbal and articulatory agility, with a total score of 26 points and higher score indicates better language function.

For the motor function, we used the Upper Extremity part of the Fugl-Meyer scale (UEFM) to evaluate the paralyzed upper limb function, which consisted 33 items and a total score of 66 points, of which higher score indicated better motor function of the upper limb. The spasticity of elbow and finger on the paralyzed side were also evaluated with the Modified Ashworth Scale (MAS), with higher values indicating more spasticity.

To explore the underlying mechanism of improvement in language function after the CC7 cross transfer surgery, we collected the resting-state functional MRI data of 14 patients out of the 16 patients before and after the surgery. To compare the changes induced by the surgery, the index of amplitude of low frequency fluctuation (ALFF) was used. The fMRI data were processed with using DPABI tookit implanted in MATLAB. The image data were preprocessed according to standard steps including slice-timing, realign, and normalized with its structural images by DARTEL. The images were smoothed with a Gaussian kernel that was 6-mm full-width at half-maximum and filtered with 0.01-0.1 Hz. The ALFF analyses were calculated in each patient.

### Statistics

For demographic data and function data at baseline, descriptive statistics was performed. For continuous variables, functional measures at three time points were analyzed by One-way Repeated Measures ANOVA, followed by Tukey’s multiple comparison test between each group. For discrete variables, multiple comparison of measurements at three time points were analyzed with Friedman test, followed by Dunn’s multiple comparisons test. Adjusted P value is reported and *\*P* < 0.05 was considered statistically significant. The ratio of change in each scale were calculated as the mean change divided by the total score of the full scale or each subtest. SPSS 22.0 software was used for calculation. Scatter plot and Pearson correlation coefficient was used to described the relationship between change of WAB-AQ and UEFM score. Paired two-sample t-test was performed in the analysis of resting-state fMRI data with the index of ALFF. The statistical threshold was set at P < 0.05 (FDR corrected).

## Results

### Study population

We identified 32 patients presented with hemiplegia combined with aphasia. Among them, 16 eligible patients were eligible and were included in this study. All 16 patients finished the follow-ups at both one-week and six-months follow-up of the WAB and motor function assessments, while 8 of them additionally finished the BDAE oral agility assessment. The flow chart is shown in Figure 1, and the patient characteristics was listed in Table 1.

**Figure 1.**
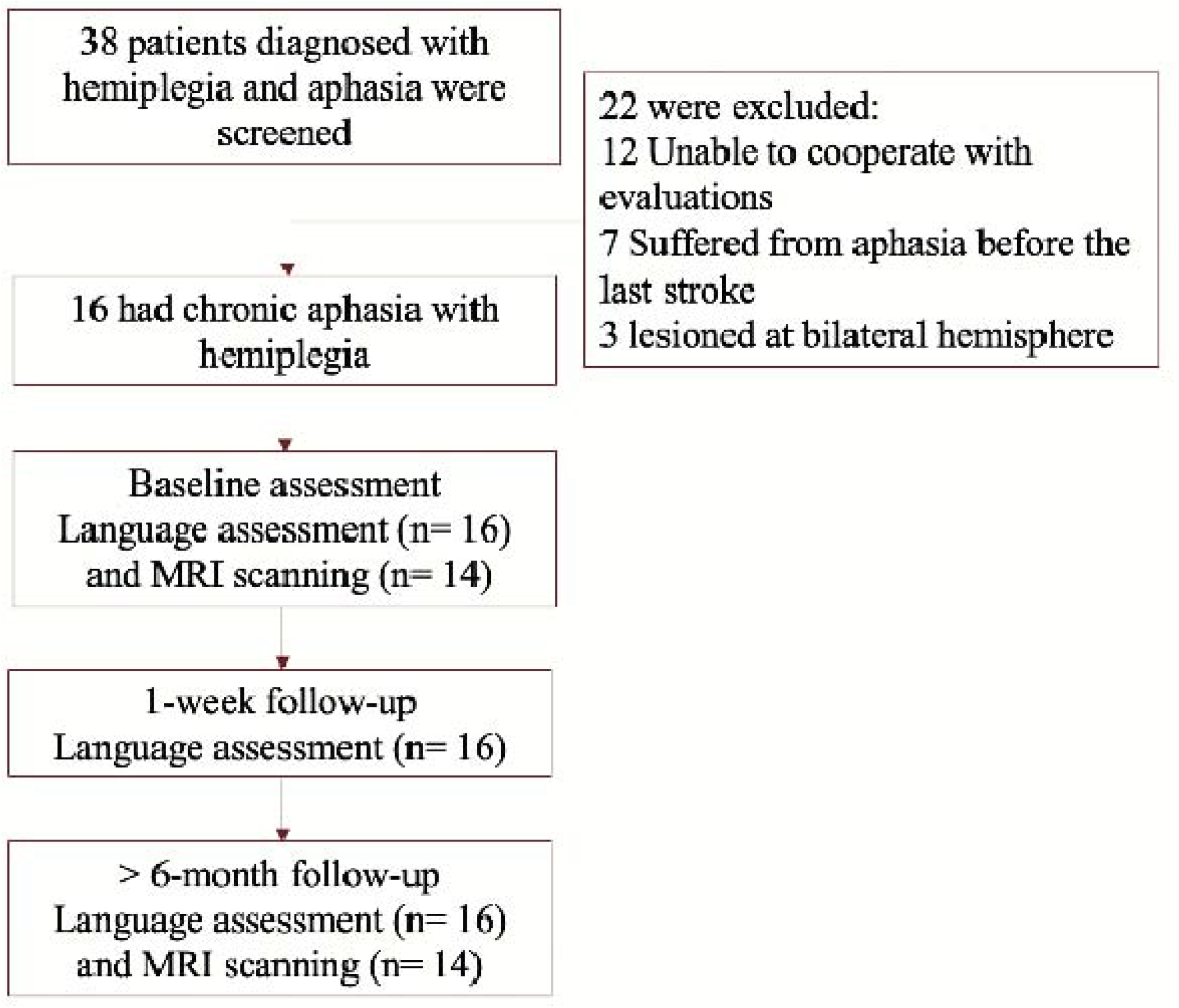
The flow chart of this study.

**Table 1.** Demographic characteristics and baseline data of the patients included.

|  | Statistics |
| --- | --- |
| **Patients included ---no. of patients** | 16 |
| **Age (Years old) --- mean±SD, range** | 45.7±15.9, range 20-71 |
| **Sex ---No. of Patients** |  |
| Male | 11 |
| Female | 5 |
| **Cause of Injury (No. of Patients)** |  |
| Ischemic stroke | 4 |
| Hemorrhagic stroke | 8 |
| Traumatic brain injury | 4 |
| **Duration of disease, years --- mean±SD, range** | 3.7±4.2, 1-19 |
| **WAB --- mean±SD, range** |  |
| Spontaneous Speech | 12.1±4.3, 2-18 |
| Comprehension | 7.2±2.0, 3.05-9.85 |
| Repetition | 7.7±2.3, 0.4-10 |
| Naming | 5.1±2.7, 0-8.4 |
| Aphasia Quotient | 63.9±21.6, 10.9-92.5 |
| **BDAE Oral Agility --- mean±SD, range\*** | 15.9±4.4, 8-22 |
| **UEFM score --- mean±SD, range** | 21.1±12.4, 3-41 |
| **MAS score ---no. of patients†** |  |
| Elbow | 3(1), 8(2), 5(3) |
| Fingers | 2(1), 9(2), 5(3) |

\*The BDAE Oral agility score was collected in 8 patients. † The MAS score was presented with the count of each score. For example, 3(1) referred to that 3 patient had 1 degree MAS score.

### Outcomes

In language function tests, the WAB-AQ showed immediate and significant increase from baseline to 1-week (mean change: 8.08, 95% CI: 0.8 to 2.6), and this change maintained stable to 6-month follow-up (mean change: 9.51, 95%CI: 6.76 to 12.27). The four subtests of the WAB scale all acquired significant increase from baseline to 1-week and 6-month follow-up, including spontaneous speech (average ratio of increase: 8.5% and 11.5%), comprehension (8.1% and 6.6%), repetition (5.3% and 7.3%) and naming (10.1% and 11.2%), respectively (see Table 2 and Figure 2). The difference between 1-week and 6-month follow-up did not meet the lowest significant level (*P*<0.05) in any of the tests or subtests, with the largest difference seen in spontaneous speech (Mean difference: 0.56, 95%CI: 0.02 to 1.14, adjusted *P*: 0.058). In the BDAE-Oral Agility evaluation in 8 patients, the change from baseline to 1-week and 6 month showed similar pattern to WAB tests, see Figure 2.

**Table 2.**
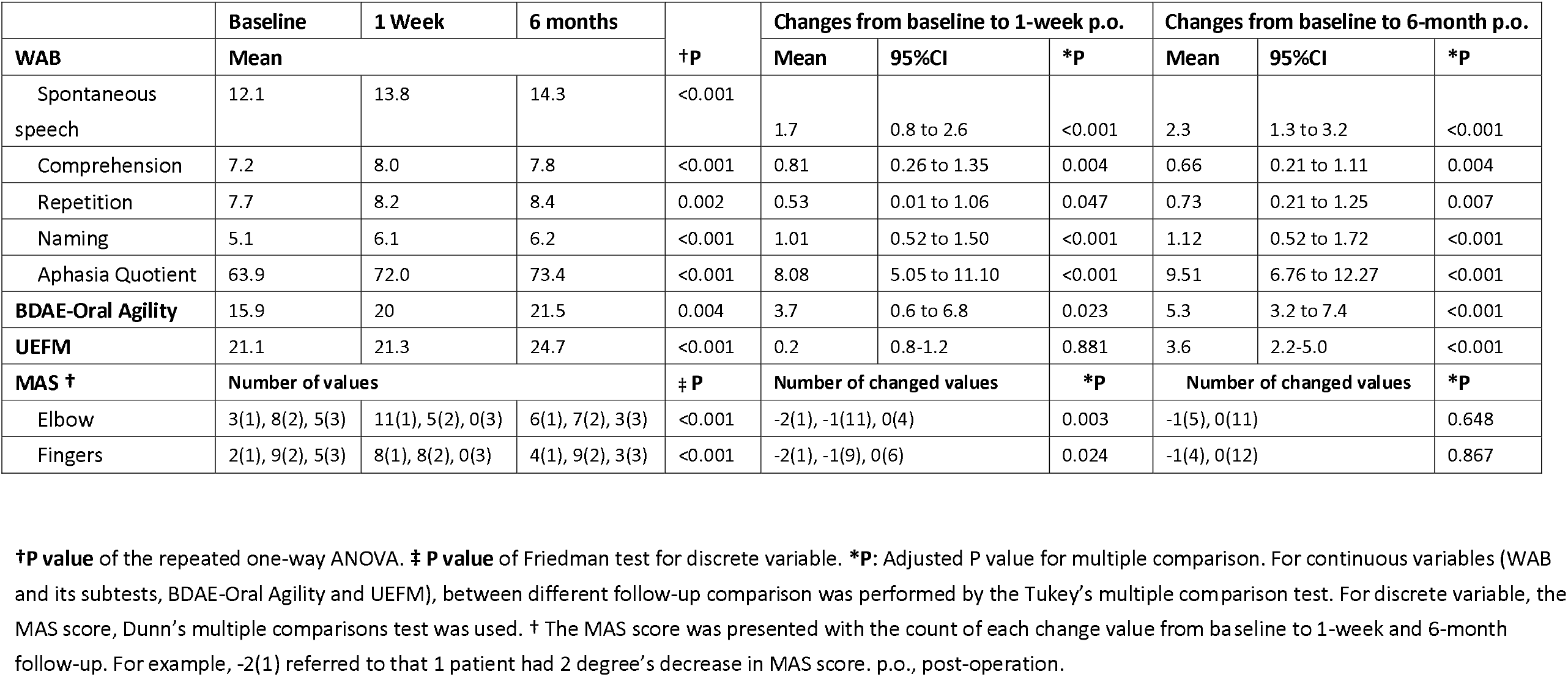
Changes of the language and motor function evaluations from baseline to each follow-up.

**Figure 2.**
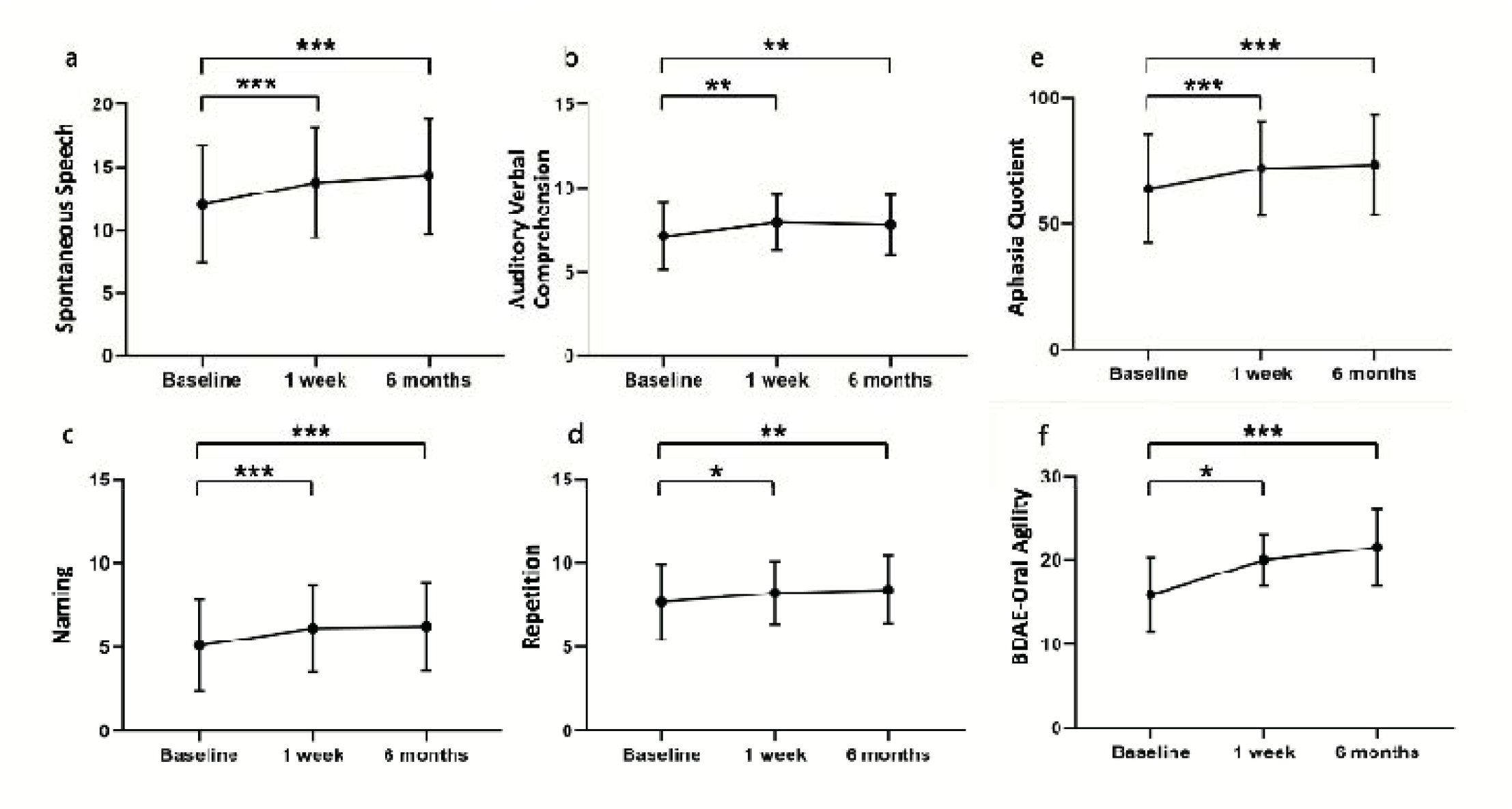
The scores of Aphasia Quotient and BDAE-oral agility tests at baseline and two follow-up points. Functional measures were analyzed by One-way Repeated Measures ANOVA, and followed by between group comparison adjusted by Tukey’s post hoc test. Adjusted *P*-values were marked, with one asterisk indicated *P* < 0.05, and two asterisks indicated *P* < 0.01, three asterisks indicated *P* < 0.001. Panel a-d: The four subsets of the WAB test in 16 patients, e: The aphasia quotient calculated by the WAB test in 16 patients. f: The BDAE oral agility test in 8 patients.

In the motor function tests, the UEFM score showed significant increase at 6-month follow-up (mean change: 3.6, 95%CI: 2.2-5.0, *P*< 0.001), but not at 1 week after the surgery (mean change: 0.2, 95%CI: 0.8-1.2, *P*=0.881). The spasticity of the elbow and fingers decreased significantly at 1 week after the surgery (*P:* 0.003 and 0.024), but partially recurred at 6-month follow-up (*P:* 0.648 and 0.867).

The scattered plot showed the relationship between the change of UEFM score and WAB-AQ score from baseline to 1-week and 6-month follow-up (Figure 3). No association between these two functional changes was shown from the plot. The Pearson correlation coefficient between the change of WAB-AQ and UEFM score from baseline to 1-week was -0.185 with a P-value of 0.493, which indicated no significant correlation between them. Similarly, the Pearson correlation coefficient between the change of WAB-AQ and UEFM score from baseline to 1-week was -0.191 with a P-value of 0.478, also show no significant correlation.

**Figure 3.**
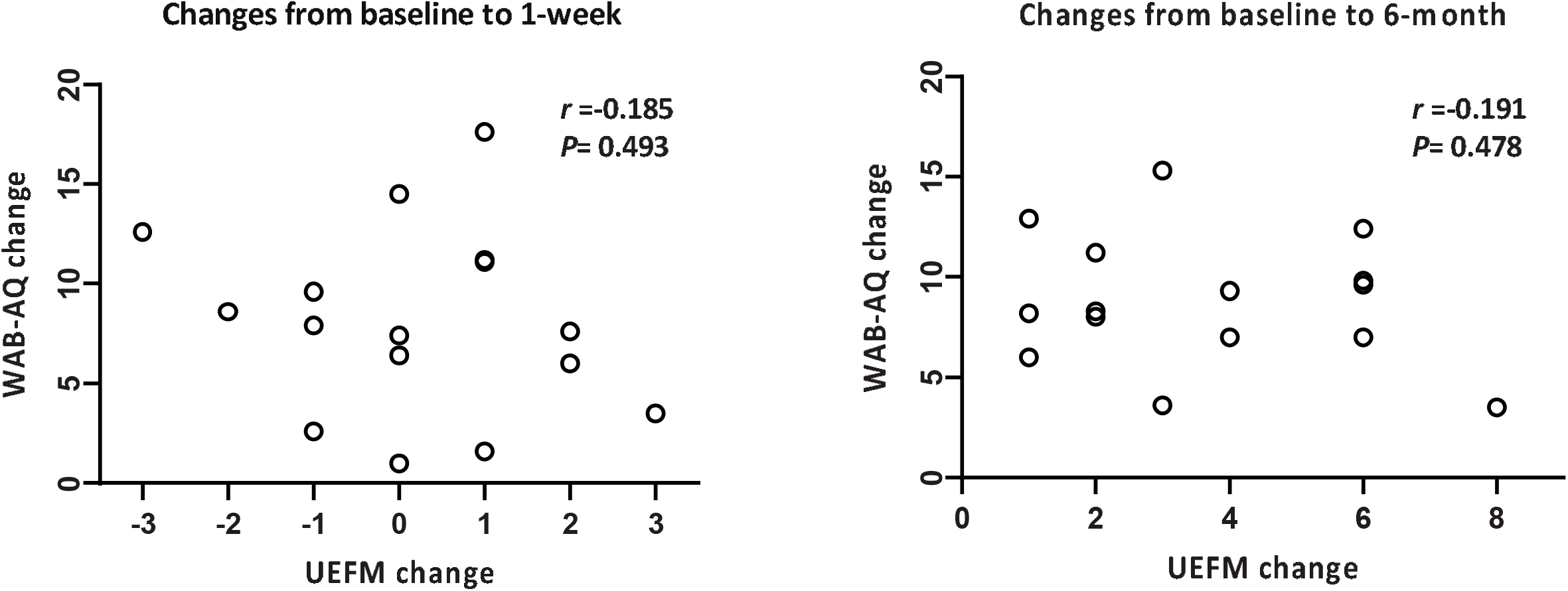
Scatter plot between the changes of WAB-AQ score and UEFM score from baseline to 1-week and to 6-month follow-up. The Pearson correlation coefficient and P value of the two change scores.

In 16 patients, 14 patients accomplished the functional MRI scanning before and after the surgery. Through paired two-sample t-test of ALFF values of two groups, six brain clusters were detected to be significantly activated after the surgery comparing to the presurgical state, including the right anterior cingulate gyrus, bilateral precentral gyrus, the left middle temporal gyrus, the orbital frontal gyrus and right gyrus rectus (Figure 4, Table 3).

**Table 3.**
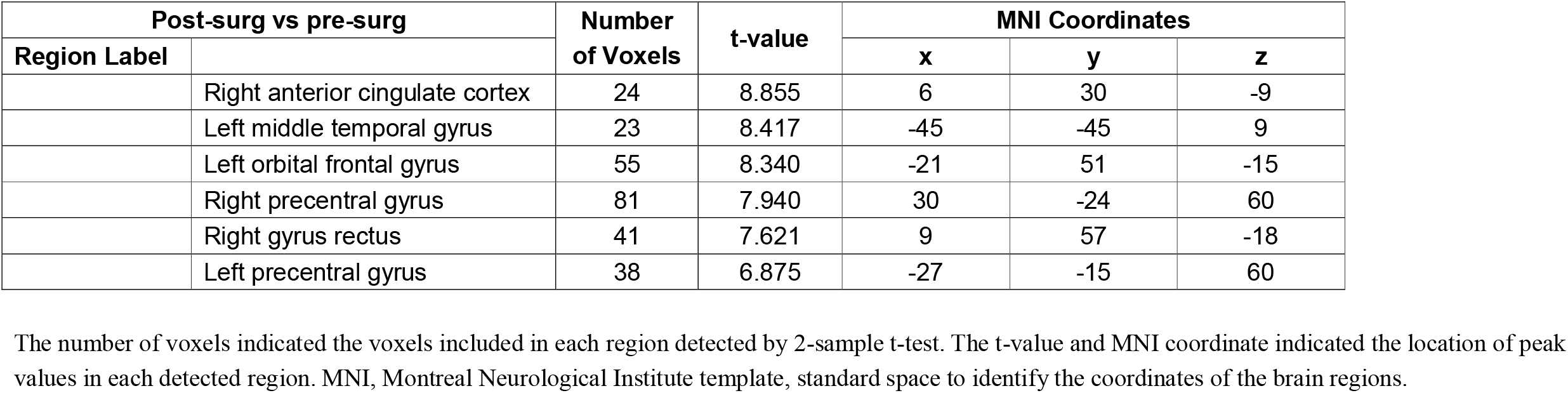
Results of 2-sample t-tests of the ALFF values between the post-surgical and pre-surgical scanning of the patients.

| Post-surg vs pre-surg |  | Number<br>of Voxels | t-value | MNI Coordinates |  |  |
| --- | --- | --- | --- | --- | --- | --- |
| Region Label |  |  |  | x | y | z |
|  | Right anterior cingulate cortex | 24 | 8.855 | 6 | 30 | -9 |
|  | Left middle temporal gyrus | 23 | 8.417 | -45 | -45 | 9 |
|  | Left orbital frontal gyrus | 55 | 8.340 | -21 | 51 | -15 |
|  | Right precentral gyrus | 81 | 7.940 | 30 | -24 | 60 |
|  | Right gyrus rectus | 41 | 7.621 | 9 | 57 | -18 |
|  | Left precentral gyrus | 38 | 6.875 | -27 | -15 | 60 |

The number of voxels indicated the voxels included in each region detected by 2-sample t-test. The t-value and MNI coordinate indicated the location of peak values in each detected region. MNI, Montreal Neurological Institute template, standard space to identify the coordinates of the brain regions.

**Figure 4.**
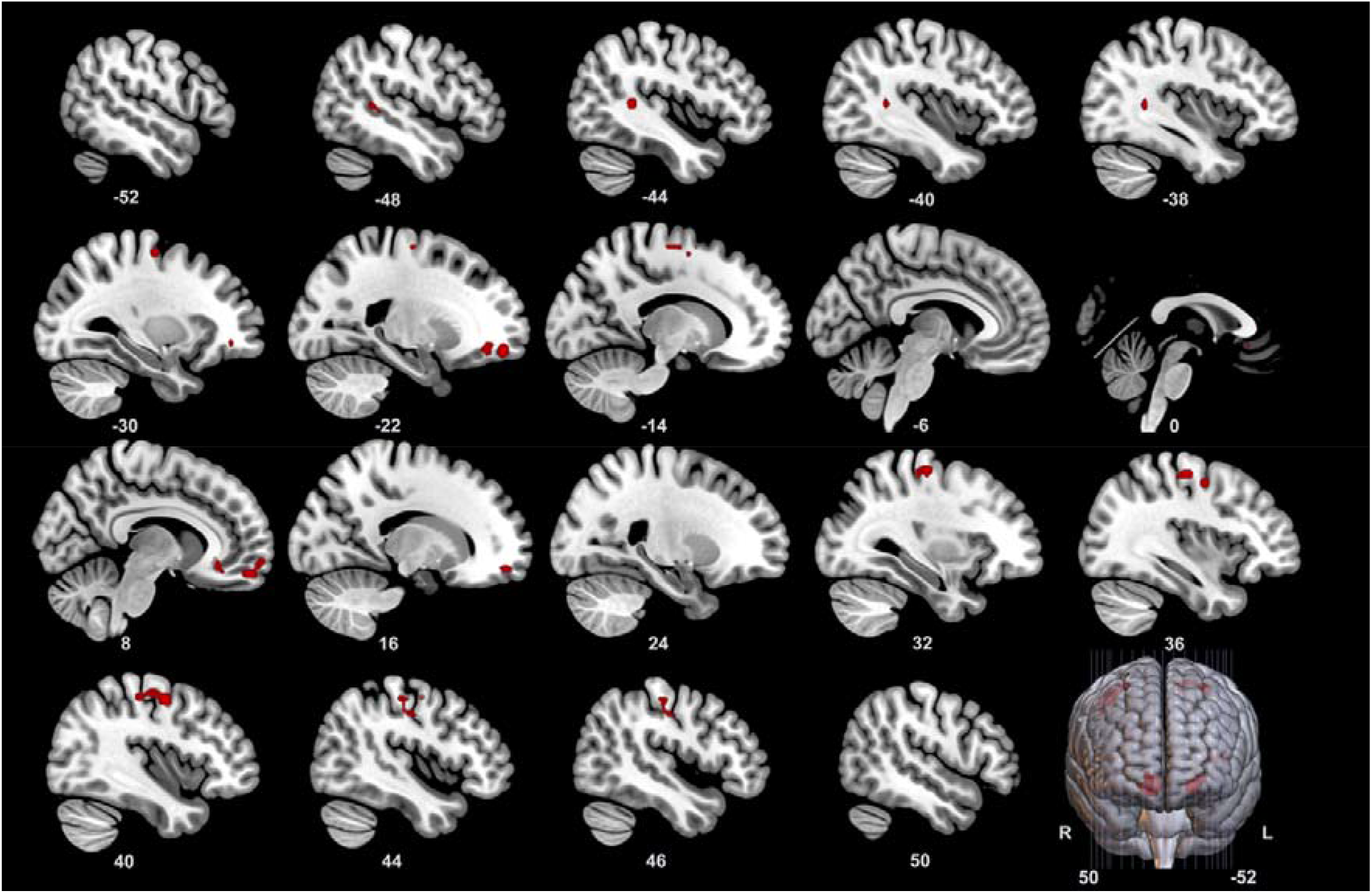
The results of paired two sample t-test of the ALFF values from the resting-state functional MRI data of the patients before and after the surgery. The results are shown as the t-value of each voxel in the figures (P<0.05, multi-comparison correction through false discovery rate). The voxels in red indicated significantly higher local activations after the surgery comparing to the data before the surgery in an average level. ALFF: amplitude of low-frequency fluctuation.

## Discussion

In this prospective cohort study, we found that the language function in hemiplegia patients with aphasia is significantly improved 1 week after CC7 cross transfer surgery, a surgery designed to improve the limb function of the paralyzed arm. This improvement is maintained to six months follow-up, although no SLT treatment are provided. Therefore, this improvement is more likely to be caused by CC7 transfer surgery, which seems to be “spontaneous” and “immediate”, with an effect size of approximately 10% of language function.

The mechanism underlying the spontaneous improvements in language function is elusive, since the CC7 cross transfer is designed to improve the motor function. In a preliminary peek of the mechanism of this phenomenon, we found that the right hemisphere was largely involved in the functional reorganization after the surgery, including the right precentral cortex, right gyrus rectus, and right anterior cingulate cortex. These regions are frequently reported to be the pivotal regions in compensation of the language function in the subacute or chronic stage^13, 17-19^. Moreover, a pivotal region that plays an important role in language production and naming function, the left middle temporal gyrus (MTG), was also significantly activated after the surgery comparing to presurgical state. The left MTG plays a crucial role in the ventral language network which is responsible for comprehension and semantically driven speech^1, 14^. These findings are in accordance with previous findings, which indicated that the improvement in language function may be related to the interhemispheric functional reorganization. While the left MTG has shown increased activity in resting-state fMRI, the involvement of bilateral hemisphere after CC7 transfer should be noticed. Regions including the right anterior cingulate cortex, right precentral gyrus, and right gyrus rectus were detected to be more active after the surgery. Among them, the right precentral gyrus showed the largest area of increased activity after the surgery. These regions may play important roles in the compensative plasticity of the language network.

From a theoretical perspective, the C7 nerve transection is more likely to be the cause of language improvement, since the functional improvement appeared at 1 week after the surgery and the nerve regeneration hasn’t begun yet. The improvement of motor function mainly appears at 1 year after the surgery in previous reports^10, 20^. Since the motor function is deeply involved in the network of language production^21, 22^, it’s more likely to be related to the spasticity decrease which also appeared at one-week. However, spasticity would recur approximately at 3 months after the surgery and sustained to over 6 months after the surgery^10^, but the improvement in language function seemed to be stable over 6 months. Further research is needed to elucidate this question.

Regardless of the mechanism of language improvement, further research should focus on how to apply this surgery to aphasia population with or without hemiplegia. The first thought would be the combination of this surgery with other rehabilitations. The commonly used interventions in treating aphasia focused on promoting the plasticity of the remaining brain ^23^, including SLT^7, 24^, noninvasive brain stimulation such as transcranial stimulation^25^. The effects of these methods were reported to be beneficial, but further improvement was still needed ^26-28^. As these interventions could vigor the compensational network for the language function, combining these methods with the CC7 nerve transfer or a simplified surgery is promising^29^. The second thought in applying this surgery would be right C7 transection alone rather than CC7 nerve cross transfer. The interpretation of the results of MAS and language function has proved the possibility that C7 transection on the right side could be the most important step for the improvement of language function, and previously reports have proved that transection of C7 nerve alone would not affect the function of the upper limb. Similar to the C8 nerve root transection, C7 transection is safe and more convenient than CC7 cross transfer surgery^30-32^. Therefore, explorative study of C7 transection in treating aphasia should be attempted through well-designed clinical trial studies.

In conclusion, CC7 cross transfer surgery could provide immediate and stable improvements in the language function in hemiplegic patients combined with chronic aphasia. These improvements were likely related to the increased activation of multiple brain areas in bilateral hemispheres, especially the right precentral gyrus, which was suggested to be a potential pivotal area of language compensation in stroke or brain injuries^1, 33, 34^. Further exploration was needed to explore more detailed mechanisms in the following studies.

### Limitation

The major limitation of this study is the study design as a preliminary observation. Randomized controlled clinical trial should be performed and more clinical evaluations as well as clinical neurophysiological studies should be added in further studies.

## Data Availability

All data produced in the present study are available upon reasonable request to the authors.

## Notes

**Interests of conflict:** The authors have no conflict of interest concerning the materials or methods used in this study or the findings specified in this article.

### Competing Interest Statement

The authors have declared no competing interest.

### Clinical Trial

ChiCTR2200060235

### Funding Statement

This study was funded by the National Natural Science Foundation of China (82021002, 81830063 and 82072539), CAMS Innovation Fund for Medical Sciences (2019-I2M-5-007), Shanghai Clinical Research Center for Aging and Medicine (19MC1910500), National key R&D program of China (2022YFC3602701), Youth Talent Project of Shanghai Municipal Health Commission (2022YQ009).

### Author Declarations

Institutional review board of Shanghai Jing An District Central Hospital gave ethical approval for this work.

